# The Novel Coronavirus, 2019-nCoV, is Highly Contagious and More Infectious Than Initially Estimated

**DOI:** 10.1101/2020.02.07.20021154

**Authors:** Steven Sanche, Yen Ting Lin, Chonggang Xu, Ethan Romero-Severson, Nick Hengartner, Ruian Ke

## Abstract

The novel coronavirus (2019-nCoV) is a recently emerged human pathogen that has spread widely since January 2020. Initially, the basic reproductive number, *R*_0_, was estimated to be 2.2 to 2.7. Here we provide a new estimate of this quantity. We collected extensive individual case reports and estimated key epidemiology parameters, including the incubation period. Integrating these estimates and high-resolution real-time human travel and infection data with mathematical models, we estimated that the number of infected individuals during early epidemic double every 2.4 days, and the *R*_0_ value is likely to be between 4.7 and 6.6. We further show that quarantine and contact tracing of symptomatic individuals alone may not be effective and early, strong control measures are needed to stop transmission of the virus.

**One-sentence summary:** By collecting and analyzing spatiotemporal data, we estimated the transmission potential for 2019-nCoV.

## Main Text

2019-nCoV is the etiological agent of the current rapidly growing outbreak originating from Wuhan, Hubei province, China (*1*). At the end of December 2019, 41 cases of ‘pneumonia of unknown etiology’ were reported by the Wuhan Municipal Health Committee (*2*). On January 1, 2020, the Huanan Seafood Wholesale Market in Wuhan, which was determined to be the epicenter of the outbreak, was closed. Seven days later, the causative agent of new disease was formally announced by China CDC as 2019-nCoV. Human-to-human transmission was later reported, i.e. infection of medical workers reported by the news and infection of individuals with no recent history of Wuhan visit (*3*). In response, China CDC upgraded the emergency response to Level 1 (the highest level) on January 15. By January 21, 2019-nCoV infection had spread to most of the other provinces. On January 23, the city of Wuhan was locked down/quarantined, all transportations into and out of the city and all mass gatherings was canceled. However, the number of confirmed cases has continued to increase exponentially since January 16 (Fig. 1A and B). On January 30, the World Health Organization (WHO) declared the outbreak a public health emergency of international concern (*4*). As of February 5, 2020, the virus outbreak lead to more than 24,000 total confirmed cases and 494 deaths, and the virus has spread to 25 countries. Initial estimates of the growth rate of the outbreak based on early case count data in Wuhan and international flight data up to mid-January were 0.1 per day (a doubling time between 6-7 days) and a basic reproductive number, *R*_0_ (defined as the average number of secondary cases an index case infects when it is introduced in a susceptible population), of 2.2 and 2.7 (*1, 5*); however, the rates of growth in the number of confirmed cases during late January (Fig. 1A and B) suggest a doubling time much shorter than 6-7 days.

**Fig. 1.**
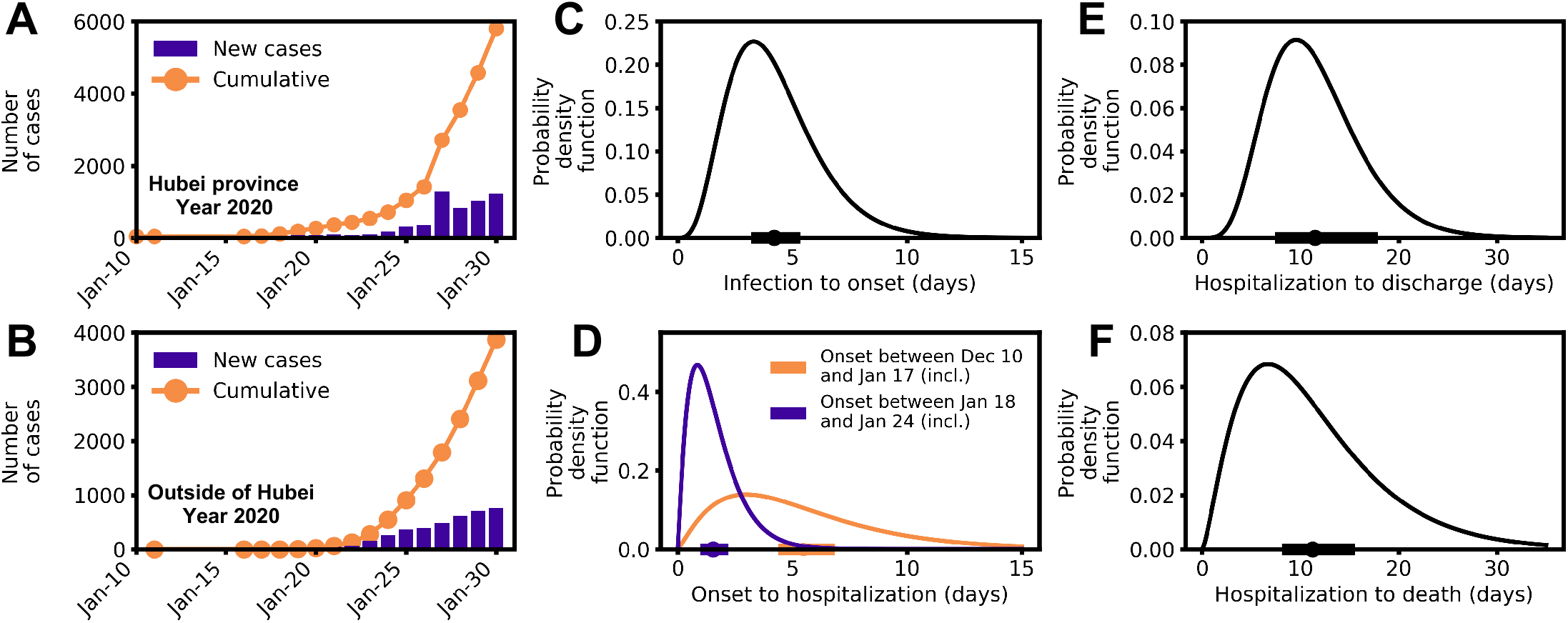
Epidemiological characteristics of early dynamics of 2019-nCoV outbreak in China. **(A-B)** Daily new and cumulative confirmed cases in Hubei province (A) and provinces other than Hubei in China (B). **(C-F)** Distributions of key epidemiological parameters, including the durations between infection and symptom onset (C), between symptom onset to hospitalization (D), between hospitalization to discharge (E) and between hospitalization to death (F). Filled circles and bars on x-axes denote the estimated mean and 95% confidence intervals.

Here, with more up-to-date and high-resolution datasets across China until the end of January, we estimated that the exponential growth rate and *R*_0_ are much higher than these previous estimates. We improved on previous estimates in three distinct ways. First, we used an expanded dataset of individual case reports based on our collection and direct translations of documents published daily from official health commissions across provinces and special municipalities in China (see Data Collection in Supplementary Materials). Second, we integrated high-resolution real-time domestic travel data in China. Third, to address the issue of potential data collection and methodological bias or incomplete control of confounding variables, we implemented two distinct modeling approaches using different sets of data. These analyses produced estimates of the exponential growth rates that are consistent with one another and higher than previous estimates.

A unique feature of our case report dataset (Table S1) is that it includes case reports of many of the first or the first few individuals who were confirmed with the virus infection in each province, where dates of departure from Wuhan were reported. All together, we collected 140 individual case reports (Table S1). These reports include demographic information including age, sex and location of hospitalization, as well as epidemiological information including potential time periods of infection, dates of symptom onset, hospitalization and case confirmation.

Using this dataset, we estimated the basic parameter distributions of durations from initial exposure to symptom onset to hospitalization to discharge or death. Our estimate of the time from initial exposure to symptom onset is 4.2 days with a 95% confidence interval (CI for short below) between 3.5 and 5.1 days (Fig. 1C). This estimated period is about 1 day shorter and has lower variance than a previous estimate (*1*). The shorter time is likely caused by the expanded temporal range of our data that includes cases occurring after broad public awareness of the disease. Patients reported in the Li et al. study (*1*) are all from Wuhan and most had symptom onset before mid-January; in our dataset, many patients had symptom onset during or after mid-January and were reported in provinces other than Hubei province (where Wuhan is the capital). The time from symptom onset to hospitalization showed evidence of time dependence (Fig. 1D and S1). Before January 18, the time from symptom onset to hospitalization was 5.5 days (CI: 4.6 to 6.6 days); whereas after January 18, the duration shortened significantly to 1.5 days only (CI: 1.2 to 1.9 days) (*p*-value <0.001 by Mann-Whitney U test). The change in the distribution coincides with the period when infected cases were first confirmed in Thailand, news reports of potential human-to-human transmission and upgrading of emergency response level to Level 1 by China CDC.The emerging consensus about the risk of 2019-nCoV likely led to significant behavior change in symptomatic people seeking more timely medical care over this period. We also found that the time from initial hospital admittance to discharge is 11.5 days (Fig. 1E; CI: 8.0 to 17.3 days) and the time from initial hospital admittance to death is 11.2 days (Fig. 1F; CI: 8.7 to 14.9 days).

Moving from empirical estimates of basic epidemiological parameters to an understanding of the actual epidemiology of 2019-nCoV requires model-based inference. We thus used mathematical models to integrate the empirical estimates with spatiotemporal domestic travel and infection data outside of Hubei province to infer the outbreak dynamics in Wuhan. Inference based on data outside of Hubei is more reliable because, as a result of the awareness of the risk of virus transmission, other provinces implemented intensive surveillance system to detect individuals with high temperatures and closely track travelers out of Wuhan using digital data to identify infected individuals (*6*) as the outbreak in Wuhan unfolded.

We collected real-time travel data during the epidemic using the Baidu® Migration server (Fig. 2A and Table S2).The server an online platform summarizing mobile phone travel data through Baidu® Huiyan [https://huiyan.baidu.com/]. Baidu® Huiyan is a widely used positioning system in China. It processes >120 billion positioning requests daily through GPS, WIFI and other means [https://huiyan.baidu.com/]. Therefore, the data represents a reliable, real-time and high-resolution source of travel patterns in China. We extracted daily travel data from Wuhan to each of the provinces. We found that in general, between 40,000 to 140,000 people in Wuhan traveled to destinations outside of Hubei province daily before the lock-down of the city on January 23, with travel peaks on January 9, 21 and 22 (Fig. 2B). Thus, it is likely that this massive flow of people from Hubei province during January facilitated the rapid dissemination of virus.

**Fig. 2.**
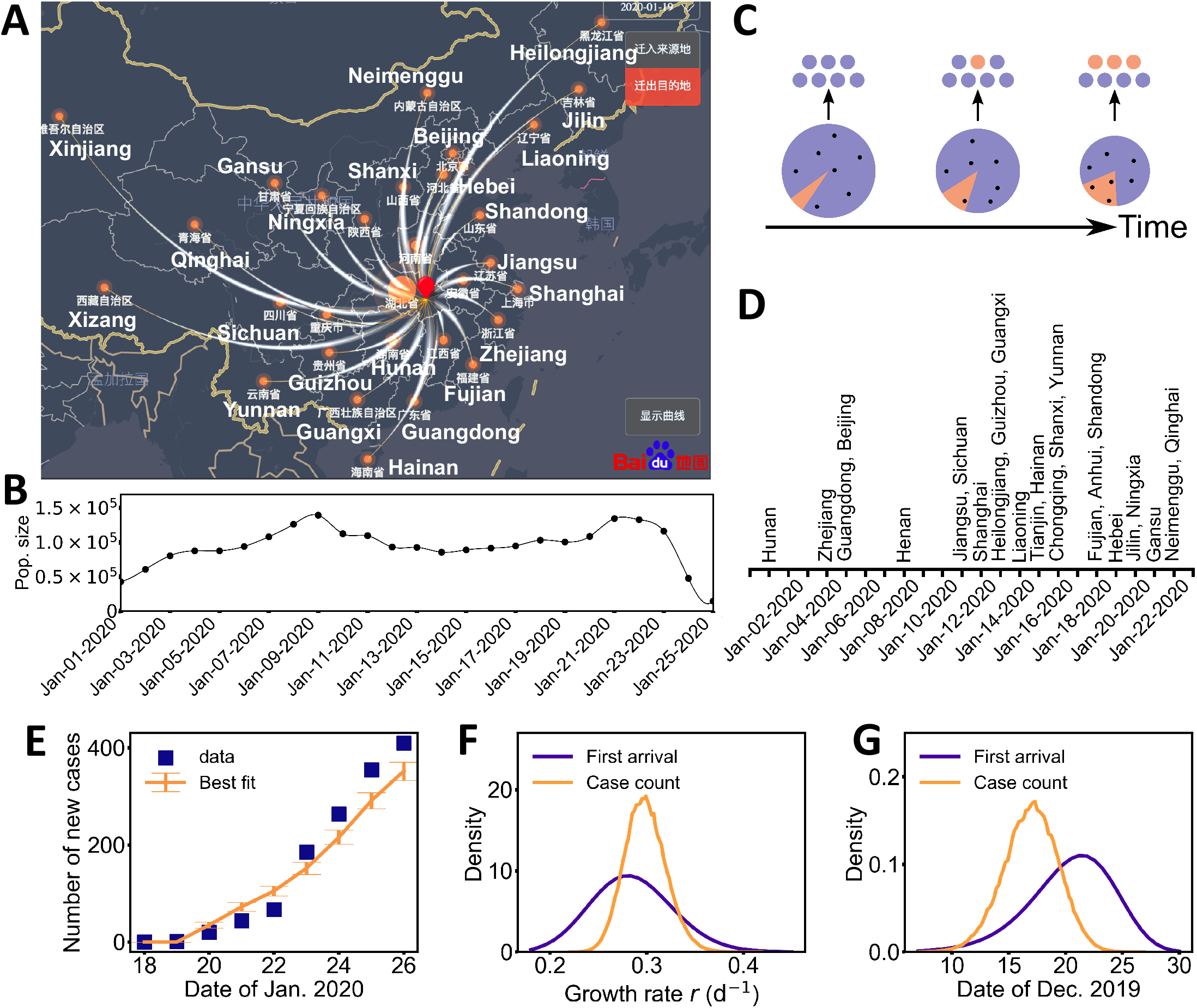
Two different approaches using high-resolution travel data reached consistent estimates of the exponential growth rate and the date of exponential growth initiation of the 2019-nCoV outbreak. **(A)** A modified snapshot of the Baidu® Migration online server interface showing the migration pattern out of Wuhan (red dot) on January 19, 2020. Thickness of curved white lines denotes the size of the traveler population to each province. The names of most of the provinces are shown in white. **(B)** Estimated daily population sizes of travelers from Wuhan, Hubei province to other provinces. **(C)** A schematic illustrating the export of infected individuals from Wuhan. Travelers (dots) are assumed to be random samples from the total population (the whole pie). Because of the growth of the infected population (orange pie) and the shrinking size of the total population in Wuhan over time, it is more and more likely infected individuals travel to other provinces (orange dots). **(D)** The dates of documented first arrivals of infected cases in 26 provinces. Names of provinces were shown vertically. **(E)** Best fit of the ‘case count’ model to daily counts of new cases (including only imported cases) in provinces other than Hubei. The standard deviations of the sample distribution are shown as the error bars. **(F and G)** The marginalized likelihoods of the growth rate *r* (F) the exponential growth initiation time (G) are consistent between the ‘first arrival’ model and the ‘case count’ model.

We integrated the travel data into our inferential models using two approaches. The rationale of the first model, the ‘first-arrival’ approach, is that an increasing fraction of people infected in Wuhan increases the likelihood that one such case is exported to the other provinces. Hence, how soon new cases are observed in other provinces can inform disease progression in Wuhan (Fig. 2C). This has similarities with earlier analyses to estimate the size of the 2019-nCoV outbreak in Wuhan based on international travel data (*5, 7, 8*), though inference based infected cases outside of China may suffer large uncertainty due to the low volume of international travel. In our model, we assumed exponential growth for the infected population *I\** in Wuhan, 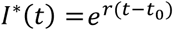, where *r* is the exponential growth rate and *t*_0_is the time of the exponential growth initiation, i.e. I^*\**^(*t*_0_)=1. Note that *t*_0_ is likely to be later than the date of the first infection event, because multiple infections may be needed before the onset of exponential growth (*9*). We used travel data to each of the provinces (Table S3) and the earliest times that an infected individual arrived at a province across a total of 26 provinces (Fig. 2D) to infer *r* and *t*_0_ (see Supplementary Materials for details). Model predictions of arrival times in the 26 provinces fitted the actual data well (Fig. S2). We estimated that the date of the beginning of an exponential growth is December 20, 2019 (CI: December 11 to 26). This suggests that human infections in early December may be due to spillovers from the animal reservoir or limited chains of transmission (*10, 11*). The growth rate of the outbreak is estimated to be 0.29 per day (CI: 0.21 to 0.37 per day), a much higher rate than two recent estimates (*1, 5*). This growth rate corresponds to a doubling time of 2.4 days. We further estimated that the total infected population size in Wuhan was approximately 4,100 (CI: 2,423 to 6,178) on January 18, which is remarkably consistent with a recently posted estimate (*7*). The estimated number of infected individuals is 18,700 (CI: 7,147, 38,663) on January 23, i.e. the date when Wuhan started lock down. We projected that without any control measure, the infected population would be approximately 233,400 (CI: 38,757 to 778,278) by the end of January (Fig. S3).

An alternative model, the ‘case count’ approach, used daily case count data between January 19 and 26 from provinces outside of Hubei to infer the initiation and the growth rate of the outbreak. We restricted the data to this period because during this time infected persons found outside of Hubei province generally reported visiting Wuhan within 14 days of becoming symptomatic, i.e. cases during that time period were indicative of the dynamics in Wuhan. We developed a meta-population model based on the classical SEIR model (*12*). We assumed a deterministic exponential growth for the infected populations in Wuhan, whereas in other provinces, we represented the trajectory of infected individuals who travelled from Wuhan using a stochastic agent-based model. The transitions of the infected individuals from symptom onset to hospitalization and then to case confirmation were assumed to follow the distributions inferred from the case report data (see Supplementary Materials for detail). Simulation of the model using best fit parameters showed that the model described the observed case counts over time well (Fig. 2E). The estimated date of exponential growth initiation is December 16, 2019 (CI: December 12 to Dec 21) and the exponential growth rate is 0.30 per day (CI: 0.26 to 0.34 per day). These estimates are consistent with estimates in the ‘first arrival’ approach (Fig. 2F and G, and Fig. S4).

We note that in both approaches, we assumed perfect detection of infected cases outside of Hubei province, i.e. the dates of first arrival and the number of case counts are accurate. This could be a reasonable assumption to make for symptomatic individuals because of the intensive surveillance implemented in China, for example, tracking individuals’ movement from digital transportation data (*6*). However, it is possible that a fraction of infected individuals, for example, individuals with mild or no symptoms (*13*), were not hospitalized, in which case we will underestimate the true size of the infected population in Wuhan. We undertook sensitivity analyses to investigate how our current estimates are affected by this issue using both approaches (see Supplementary Materials for detail). We found that if a proportion of cases remained undetected, the time of exponential initiation would be earlier than December 20, translating into a larger population of infected individuals in January, but the estimation of the growth rate remained the same. Overall, the convergence of the estimates of the exponential growth rate from the two approaches emphasizes the robustness of our estimates to model-dependent assumptions.

Our estimated outbreak growth rate is significantly higher than two recent reports where the growth rate was estimated to be 0.1 per day (*1, 5*).This estimate were based on early case counts from Wuhan (*1*) or international air travel data (*5*). However, these data suffer from important limitations. The reported case counts in Wuhan during early outbreak are likely to be underreported because of many factors, and because of the low numbers of individuals traveling abroad compared to the total population size in Wuhan, inference of the infected population size and outbreak growth rate from infected cases outside of China suffers from large uncertainty (*7, 8*). Our estimated exponential growth rate, 0.29/day (a doubling time of 2.4 days) is consistent the rapidly growing outbreak during late January (Fig. 1A).

Using the exponential growth rate, we next estimated the range of the basic reproductive number, *R*_*0*_. It has been shown that this estimation depends on the distributions of the latent period (defined as the period between the times when an individual infected and become infectious) and the infectious period (*14*). For both periods, we assumed a gamma distribution and varied the mean and the shape parameter of the gamma distributions in a large range to reflect the uncertainties in these distributions (see Supplementary Materials). It is not clear when an individual becomes infectious; thus, we considered two scenarios: 1) the latent period is the same as the incubation period, and 2) the latent period is 2 days shorter than the incubation period, i.e. individuals start to transmit 2 days before symptom onset. Integrating uncertainties in the exponential growth rate estimated from the ‘first arrival’ approach and the uncertainties in the duration of latent and infectious periods, we estimated the values of *R*_0_ to be 6.3 (CI: 3.3 to 11.3) and 4.7 (CI: 2.8 to 7.6), for the first and second scenarios, respectively (Fig. 3A). When using the estimates from the ‘case count’ approach, we estimated slightly higher *R*_*0*_ values of 6.6 (CI: 4.0 to 10.5) and 4.9 (CI: 3.3 to 7.2), for the first and second scenarios, respectively (Fig. S5). Overall, we report *R*_*0*_ values are likely be between 4.7 and 6.6 with a CI between 2.8 to 11.3. We argue that the high *R*_*0*_ and a relatively short incubation period lead to the extremely rapid growth of the of 2019-nCoV outbreak as compared to the 2003 SARS epidemic where *R*_*0*_ was estimated to be between 2.2 to 3.6 (*15, 16*).

**Fig. 3.**
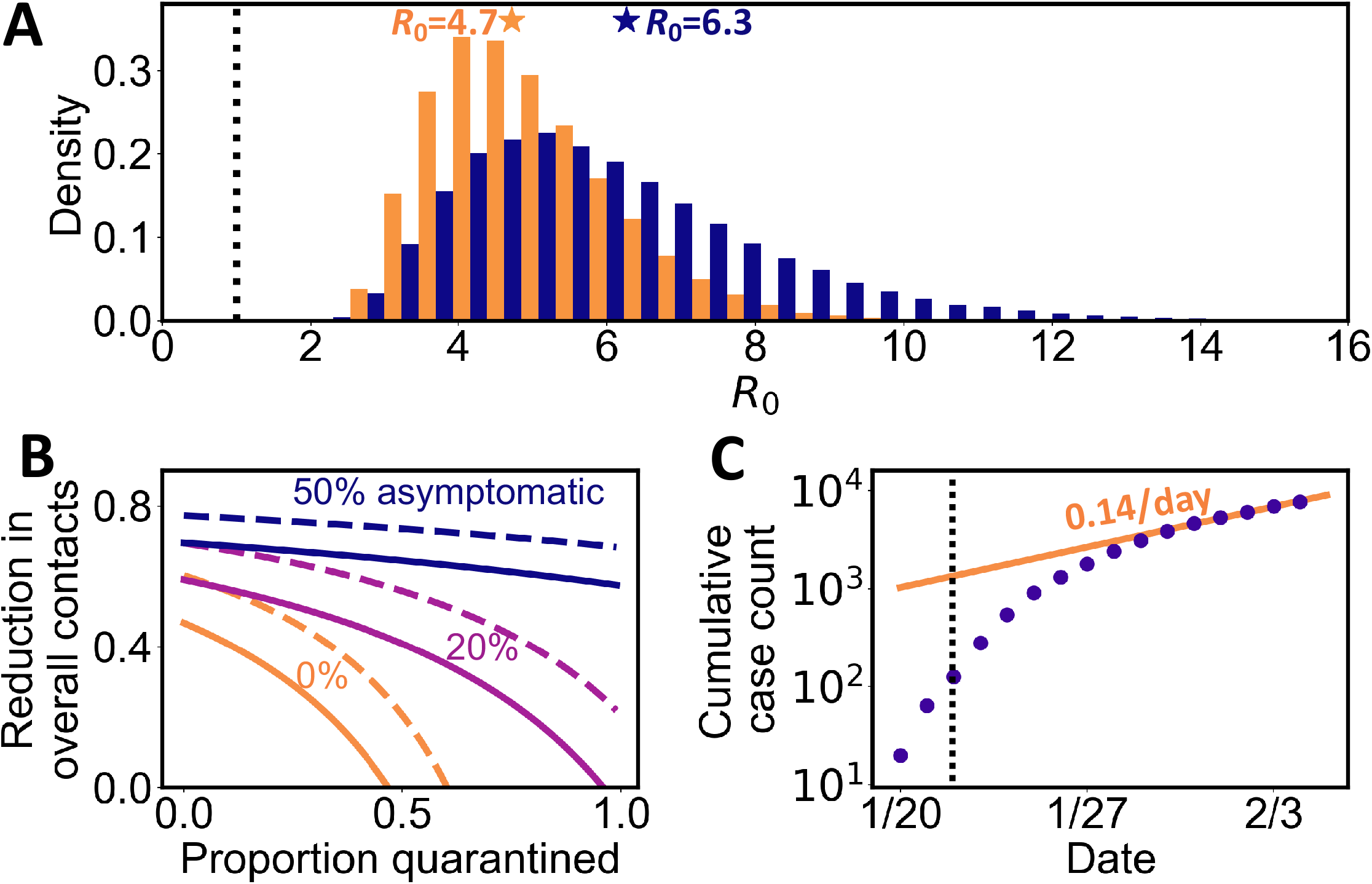
Estimation of the basic reproductive number, *R*_0_, and the impact of control measures. **(A)** Histograms and the means (stars) of estimated *R*_0_ assuming individuals become infectious at symptom onset (blue) or 2 days before symptom onset (orange). The dotted line denotes *R*_0_=1. **(B)** The levels of minimum efforts (lines) of intervention strategies needed to control the virus spread. Strategies considered are quarantine of symptomatic individuals and individuals who had contacts with them (x-axis) and population-level efforts to reduce overall contact rates (y-axis). Different colored lines denote different assumptions of the fraction of asymptomatic individuals in the infected population. Solid and dashed lines correspond to *R*_0_=4.7 and 6.3 (i.e. the estimated means of *R*_0_), respectively. **(C)** The cumulative number of cases outside of Hubei province in late January 2020. The growth rate decreased to 0.14 per day since January 30. The dashed black line shows January 23 when Wuhan is locked down.

The high *R*_*0*_ values we estimated have important implications for disease control. For example, basic theory predicts that the force of infection has to be reduced by 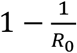to guarantee extinction of the disease. At *R*_0_ = 2 2this fraction is only 55%, but at *R*_0_= 6.7this fraction rises to 85%. To translate this into meaningful predictions, we use the framework proposed by Lipsitch et al (*16*) with the parameters we estimated for 2019-nCoV. Importantly, given the recent report of transmission of the virus from asymptomatic individuals (*13*), we considered the existence of a fraction of infected individuals who is asymptomatic and can transmit the virus (see Supplementary Materials). Results show that if as low as 20% of infected persons are asymptomatic and can transmit the virus, then even 95% quarantine efficacy will not be able to contain the virus (Fig. 3B). Given the rapid rate of spread, the sensitivity of control effort effectiveness to asymptomatic infections and the potential of transmission before symptom onset, we need to be aware of the difficulty of controlling 2019-nCoV once it establishes in a new population (*17*). Future field, laboratory and modeling studies aimed to address the unknowns, such as the fraction of asymptomatic individuals, the time when individuals become infectious and the existence of superspreaders are needed to accurately predict the impact of various control strategies (*9, 17*).

Fortunately, we see evidence that control efforts have a measurable effect on the rate of spread. Since January 23, Wuhan and other cities in Hubei province implemented vigorous control measures, such as closing down transportation and mass gatherings in the city; whereas, other provinces also escalated the public health alert level and implemented strong control measures. We noted that the growth rate of the daily number of new cases in provinces outside of Hubei slowed down gradually since late January (Fig. 3B). Due to the closure of Wuhan (and other cities in Hubei), the number of cases reported in other provinces during this period shall start to track local infection dynamics rather than imports from Wuhan. We estimated that the exponential growth rate is decreased to 0.14 per day (CI: 0.12 to 0.15 per day) since January 30. Based on this growth rate and an *R*_0_ between 4.7 to 6.6 before the control measures, a calculation following the formula in Ref. (*14*) suggested that a growth rate decreasing from 0.29 per day to 0.14 per day translates to a 50%-59% decrease in *R*_*0*_ to between 2.3 to 3.0. This is in agreement with previous estimates of the impact of effective social distancing during 1918 influenza pandemic (*18*). Thus, the reduction in growth rate may reflect the impact of vigorous control measures implemented and individual behavior changes in China during the course of the outbreak.

The 2019-nCoV epidemic is still rapidly growing and spread to more than 20 countries as of February 5, 2020. Here, we estimated the growth rate of the early outbreak in Wuhan to be 0.29 per day (a doubling time of 2.4 days), and the reproductive number, *R*_0_, to be between 4.7 to 6.6 (CI: 2.8 to 11.3). Among many factors, the Lunar New Year Travel rush in early and mid-January 2020 may or may not play a role in the high outbreak growth rate, although SARS epidemic also overlapped with the Lunar New Year Travel rush. How contiguous the 2019-nCoV is in other countries remains to be seen. If the value of *R*_0_ is as high in other countries, our results suggest that active and strong population-wide social distancing efforts, such as closing down transportation system, schools, discouraging travel, etc., might be needed to reduce the overall contacts to contain the spread of the virus.

## Data Availability

All data is available in the main text or the supplementary materials.

## Acknowledgments

We would like to thank Alan Perelson and Christiaan van Dorp for suggestions and critical reading of the manuscript and Weili Yin for help with collecting and translating documents from provincial health commission websites.

## Author contributions

RK and NH conceived the project; RK collected data; SS, YTL, CX and RK performed analyses; SS, YTL, ERS, NH and RK wrote and edited the manuscript.

## Data and materials availability

all data is available in the main text or the supplementary materials.

## Supplementary Materials

Supplementary Text

Figures S1-S5

Tables S1-S3

